# Synthetic Data Generation in Healthcare: A Scoping Review of reviews on domains, motivations, and future applications

**DOI:** 10.1101/2024.08.09.24311338

**Authors:** Miguel Rujas, Rodrigo Martín Gómez del Moral Herranz, Giuseppe Fico, Beatriz Merino-Barbancho

## Abstract

The development of Artificial Intelligence (AI) in the healthcare sector is generating a great impact. However, one of the primary challenges for the implementation of this technology is the access to high-quality data due to issues in data collection and regulatory constraints, for which synthetic data is an emerging alternative. This Scoping review analyses reviews from the past 10 years from three different databases (i.e., PubMed, Scopus, and Web of Science) to identify the healthcare domains where synthetic data are currently generated, the motivations behind their creation, their future uses, limitations, and types of data. A total of 13 main domains were identified, with Oncology, Neurology, and Cardiology being the most frequently mentioned. Five types of motivations and three principal future uses were also identified. Furthermore, it was found that the predominant type of data generated is unstructured, particularly images. Finally, several future work directions were suggested, including exploring new domains and less commonly used data types (e.g., video and text), and developing an evaluation benchmark and standard generative models for specific domains.

## 1. Introduction

The development and application of Artificial Intelligence (AI) in contemporary society is growing rapidly [1]. This technology has proven its capacity to revolutionize and influence the progress of various industries, including agriculture, transportation, and education [2]. Nevertheless, one of the most impactful areas for AI is the healthcare sector. In recent years, AI has demonstrated the potential to assist healthcare professionals in enhancing the diagnosis, treatment, and monitoring of different diseases [3]. The expected impact of AI extends beyond clinical improvements to significant economic benefits, with projected savings between 200 and 300 billion dollars only in the United States [4].

The implementation of AI in healthcare faces numerous constraints and barriers, including ethical, technological, regulatory, liability, personnel, patient safety and social issues [5]. A crucial factor in this context is the availability and quality of data, which can accelerate the implementation process by addressing some of these barriers and encouraging open scientific research. This emphasis on data aligns with a broader shift within the AI paradigm from a model-centric to a data-centric approach, where improving data access and quality is paramount to developing better AI systems [6].

In healthcare, access to high-quality data is particularly challenging due to the difficulties involved in data collection, such as the low prevalence of rare diseases, the critical conditions of some patients and the added burden for medical professionals [7]. In addition, privacy issues pose major obstacles due to the sensitive nature of health data and their potential misuse. Various techniques, such as federated learning and advanced encryption methods, are used to address these privacy concerns. However, an increasingly popular alternative is the generation of synthetic data, which can mitigate some privacy, access, intellectual property and regulatory issues while still providing valuable information [8].

Synthetic data can be defined as an artificial re-expression of real data through statistical processes, designed to mitigate privacy concerns and promote broad dissemination and open science [9]. The main goal of synthetic data is to provide a data resource that can be used for a variety of applications, such as testing and training machine learning models, while avoiding some of the risks and limitations associated with real data. Another advantage of synthetic data is its potential to improve fairness in artificial intelligence models. Synthetic datasets can be manipulated to better represent populations rather than simply reflecting the current state of the world. This means they can be designed to avoid racial or gender discrimination, helping to mitigate biases that might otherwise be present in real-world data.

Despite their advantages, synthetic data also present some challenges, particularly in terms of monitoring the results. Ensuring the accuracy and consistency of results from synthetic data can be complex, especially with complex datasets. The quality of synthetic data also depends heavily on the quality of the original data and the data generation model used. If the original data contain biases, these biases may be reflected in the synthetic data, potentially compromising its unbiasedness and usefulness. In addition, efforts to manipulate datasets to create fair synthetic data may inadvertently lead to inaccuracies, as overly sanitised data may not accurately reflect real-world conditions. These challenges indicate the importance of scrutiny and rigorous validation of synthetic data quality, also assuring the explainability for AI applications. In addition, all these considerations must be in line with compliance with emerging regulations such as the EU AI Act, first-ever legal framework on AI, which includes provisions for the use of synthetic data under *Art*.*10 (Data and data governance)*, especially for training, validation and testing data sets in a high-risk sector such as healthcare.

As the field continues to evolve, there is a need to explore and understand the specific health domains in which synthetic data are generated and use cases addressing underrepresented data types like device, image, and genomic data. This comprehension may help to identify best practices, address potential bottlenecks and maximise the benefits of synthetic data in advancing health innovations.

### 1.1. Related work

Gonzales et al. [6] conducted a narrative review exploring the potential applications of synthetic data in healthcare. This review highlights the importance of synthetic data in bridging the gap in data accessibility, addressing privacy concerns, and enabling innovative applications. The authors identified seven potential use cases for synthetic data in healthcare, including simulation and prediction research, hypothesis and algorithm testing, epidemiology, health IT development, education and training, public release of datasets, and data linking. They also discussed the limitations and challenges of using synthetic data (e.g., data leakage risks).

Hernandez et al. [9] performed a systematic review focusing on the technological aspects of tabular data generation, with special emphasis on privacy-preserving techniques. This review analyses the different approaches for generating tabular synthetic data, especially utilizing generative adversarial networks (GANs) (e.g., Medical GAN or Supervised GAN), and for evaluating key aspects, including resemblance, utility, and privacy. They also discuss the challenges associated with maintaining data privacy while ensuring utility.

Murtaza et al. [10] presented a state-of-the-art overview of synthetic data generation in healthcare, categorising the approaches into three main types: Knowledge-Driven, Data-Driven, and Hybrid. This review defined the essential attributes of synthetic data, including realism and privacy, and examined different methods and metrics used for generating and evaluating synthetic data. The authors provided insights into the current technologies used for synthetic data generation, along with a discussion on the potential future directions of synthetic data in healthcare.

While these reviews focus on the techniques and strategies for generating synthetic data and evaluating these models, there is a gap in the literature regarding the specific healthcare domains and subdomains where synthetic data is being generated. Additionally, the motivations driving the creation of synthetic data and its intended future applications within these areas have not been explored. Understanding these aspects is important for identifying trends, challenges, and opportunities in the application of synthetic data in healthcare.

As this is a relatively new area where research is being conducted to understand how synthetic data in health is generated and under what assumptions and domains, we propose that there is value in specifically examining the reviews already published in this area as it robustly aggregates existing knowledge to date. In doing so, we hope to identify domains of health where synthetic data are already being applied, the motivations underlying the creation of these synthetic data, and the applications. In this regard, a scoping review approach is suited for this purpose, as it provides a comprehensive mapping of the existing literature, identifying these characteristics and emerging trends. Therefore, by analysing reviews from the last decade that focus on specific health domains (e.g., cardiology, oncology), this scoping review aims to provide an overview of the existing reviews in the literature. The primary research question guiding this review is: “*In which main healthcare domains and subdomains is synthetic data being generated, what motivations drive its creation within these areas, and what are the envisioned future applications of this data?*”. In this scoping review of reviews, we mapped the existing literature describing the approaches under which synthetic data in healthcare is being generated and applied.

## 2. Materials and Methods

The methodology employed for this Scoping review adheres to the guidelines established in the *“Preferred Reporting Items for Systematic reviews and Meta-Analyses extension for Scoping Reviews (PRISMA-ScR)”* [11]. The strategy followed included defining the research question, identifying and selecting relevant studies, and charting and reporting the findings.

### 2.1. Search strategy

The literature search was performed on April 25, 2024, across PubMed, Web of Science, and Scopus, utilizing key terms associated with synthetic data and healthcare (e.g., “*synthe\**”, “*record*” or “*health\**”). The queries were further refined by applying various filters available within each search engine. The complete search strategy is detailed in Table 1.

**TABLE 1.**
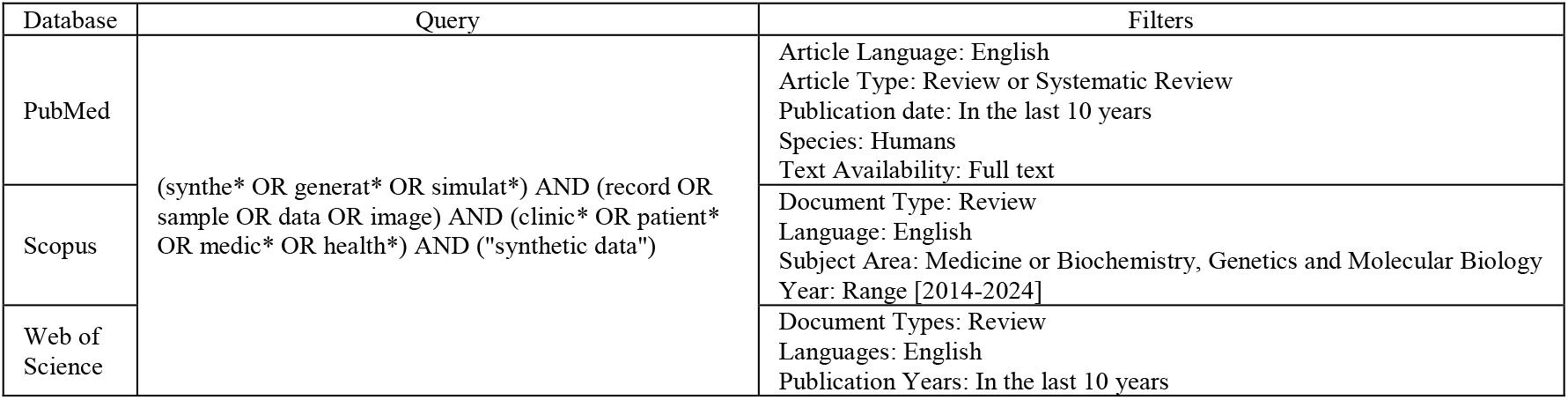
Search strategy.

### 2.2. Inclusion and exclusion criteria

The inclusion and exclusion criteria were established following the JBI methods for a Scoping Review [12]. Articles were included in the review if they met the following conditions: (1) the study involved human subjects; (2) the publication was a review or systematic review because they already analyse the limitations and gaps in the primary literature so that by including only reviews, less researched areas and opportunities for future research can be easily identified; and (3) the study was published between 2014 and 2024. Articles were excluded based on the following criteria: (1) the study was not published in English; (2) the focus of the article was not related to health; (3) the study did not fall within the scope of the research questions; (4) the study involved animal subjects; (5) the article was later than 10 years; and (6) the publication type was a book, paper, clinical trial, meta-analysis, or randomized controlled trial.

### 2.3. Search and screening process

Following the article search and extraction process, duplicate articles were removed. Initially, two independent authors (M.R. and B.M.) reviewed the titles and abstracts to identify articles suitable for inclusion in the study. Any disagreements regarding the inclusion or exclusion of an article were resolved by a third reviewer (R.M.G.). Subsequently, the full-text articles were retrieved for a second review conducted by two reviewers (M.R. and R.M.G.). Data were then extracted from the articles that met the review criteria, including: authors, main healthcare domain and subdomain, motivations for creating synthetic data, future uses of the synthetic data, type of data generated, limitations identified by the authors related to the generation of synthetic data, and type of review. If there were any uncertainties during the data extraction process, the referenced article within the review study was thoroughly examined to extract the necessary information.

### 2.4. Data charting, appraisal and synthesis of results

The various extracted characteristics were documented using an Excel spreadsheet. For the fields of motivations, future uses, type of data, and limitations, the data were represented as enumerated lists, as multiple items from each category might be present within a single study. Additionally, within the data type category, a two-level classification was performed: initially determining whether the data was structured or unstructured, and subsequently specifying the exact type of data (e.g., images or text). After extraction, the data was analysed field by field using counters and groupings to better understand and report the different trends and patterns in this topic.

## 3. Results

The study results are organized as follows: first, the numerical outcomes of the search and screening process will be displayed using a PRISMA chart, providing clarity on the process and aiding future similar reviews. Next, a table with the information extracted from the articles will be presented. Finally, the specific results for each field of extracted information will be detailed.

### 3.1. Search and screening results

The PRISMA chart illustrating the search and screening process is shown in Figure 1.

**Figure 1.**
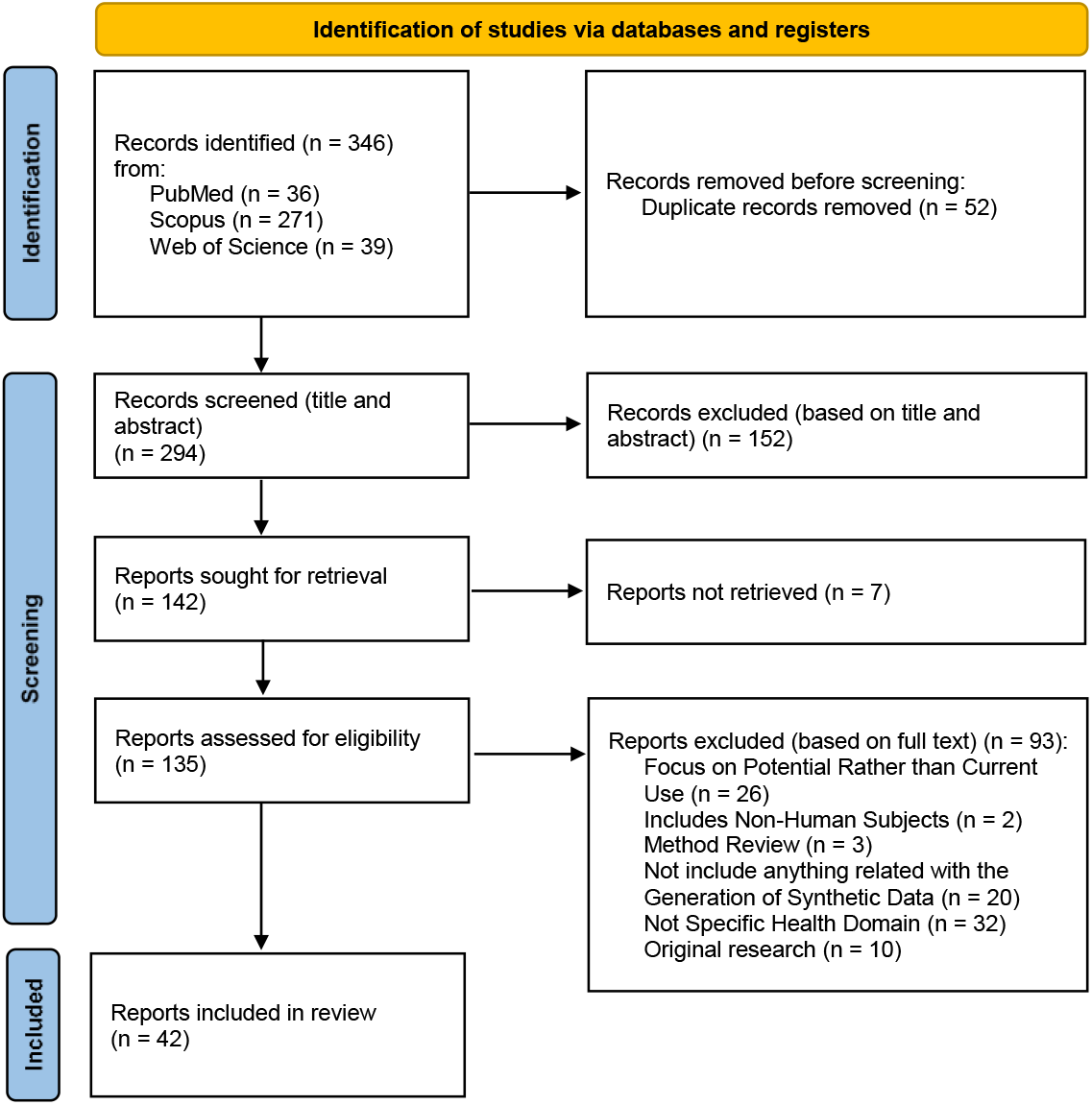
PRISMA Chart of the Scoping review

Initially, 346 articles were retrieved from three search engines: 271 from Scopus, 39 from Web of Science, and 36 from PubMed. After removing duplicates, the titles and abstracts of 294 articles were reviewed. Of these, 142 articles passed the initial screening; however, the full text of 7 articles could not be retrieved. Following the full-text review, 42 articles were included for data extraction, while 93 were excluded for the following reasons: they did not address a specific healthcare domain (e.g., cardiology or oncology) but rather a broader topic (e.g., medical imaging or precision medicine) (n = 32); they discussed the potential for synthetic data generation rather than its current application (n = 26); they did not include anything related to synthetic data generation (n = 20); they were original research articles (n = 10); they were Method reviews that compared different synthetic data generation methods without providing real evidence of synthetic data generation in the domain beyond the study itself (n = 3); or they included non-human subjects (n = 2);

### 3.2. Data extraction results

The information extracted from the various articles is summarized in Table 2.

**TABLE 2.** Summary of Extracted Information from Included Articles.

| Author | Main Healthcare Domain | Healthcare subdomain | Motivations | Future Use | Data Types | Limitations of each article | Review Type |
| --- | --- | --- | --- | --- | --- | --- | --- |
| Mirikharaji et al. [13] | Dermatology | Skin cancer | Lack of large annotated databases available | Improving the performance of AI Models | Unstructured: Images | No limitations identified in this paper | Survey |
| Monachino et al. [14] | Cardiology | Electrocardiogram | 1. Data scarcity due to limited samples (low disease prevalence)<br>2. High Unbalanced datasets<br>3. Missing data labels<br>4. Low data accessibility due to regulatory frameworks, ethics and privacy (GDPR) | 1. Developing AI Models<br>2. Enabling secondary use of data and data sharing | Structured data: Time Series | 1. Lack of a standard evaluation benchmark<br>2. If a dataset is too small and heterogeneous, it may not be possible to learn the underlying data properties effectively<br>3. GAN technical limitations | Not defined |
| Chandrabhatha et al. [15] | Ophthalmology | Intraocular cancers | Data scarcity due to limited samples (low disease prevalence) | Improving the performance of AI Models | Unstructured: Images | No limitations identified in this paper | Narrative review |
| Osuala et al. [16] | Oncology | Cancer imaging | 1. Data scarcity due to limited samples (low disease prevalence)<br>2. High heterogeneity between tumours<br>3. Low data quality<br>4. Missing data annotation<br>5. High Unbalanced datasets<br>6. Difficulty in collecting large consented datasets of highly vulnerable patients under demanding care plans (Data sharing and privacy)<br>7. Improving cancer detection, diagnosis, tumour profiling, treatment planning and monitoring | 1. Conducting analysis and studies<br>2. Developing AI Models<br>3. Improving the generalizability and interpretability of AI Models | Unstructured: Images | 1. Synthetic data generators reproduce the bias present in the training data<br>2. Variability of the synthetic data generators is limited to the training data<br>3. GAN technical limitations<br>4. Lack of a standard evaluation benchmark | Systematic review and meta-analysis |
| Skandarani et al. [17] | Cardiology | Applications of GAN in Cardiology | 1. Avoid time consuming data collection and labelling<br>2. Improving the quality of images<br>3. Handling missing data<br>4. Avoid data privacy challenges<br>5. Disease simulation and surgery planning | 1. Conducting analysis and studies<br>2. Developing AI Models<br>3. Enabling secondary use of data and data sharing<br>4. Medical education | 1. Unstructured: Images<br>2. Structured data: Time Series<br>3. Structured data: Cross-Sectional | 1. GAN technical limitations<br>2. Lack of a standard evaluation benchmark<br>3. Synthetic data generators reproduce the bias present in the training data | Not defined |
| Halfpenny and Baxter [18] | Ophthalmology | Medical information sharing | 1. Avoid data privacy challenges<br>2. De-identification | Developing AI Models | Unstructured: Images | No limitations identified in this paper | Not defined |
| Chen et al. [19] | Neurology | Cerebrovascular diseases | 1. Data scarcity due to limited samples<br>2. High Unbalanced datasets | Developing AI Models | Structured data: Cross-Sectional | ADASYN's performance is limited with high imbalanced datasets | Systematic review |
| Metzcar et al. [20] | Oncology | Mathematical Oncology | Avoid data privacy challenges | 1. Data sharing<br>2. Developing Large-scale Models | Unstructured: Images | Synthetic data generators reproduce the bias present in the training data | Not defined |
| Lou et al. [21] | Neurology | Facial nerve function assessment | Building a benchmark database for evaluation purposes | 1. Developing AI Models<br>2. Testing AI Models | Unstructured: Images | No limitations identified in this paper | Systematic review |
| Mohsen et al. [22] | Endocrinology | Diabetes | High Unbalanced datasets | Not specified | Structured data: Cross-Sectional | No limitations identified in this paper | Scoping review |
| Bai et al. [23] | Oncology | Breast cancer detection | 1. Improving the quality of the data<br>2. Enable comparison between different methods<br>3. Improving ML Systems | Developing AI Models | Unstructured: Images | Synthetic data generators reproduce the bias present in the training data | Not defined |
| Lipkova et al. [24] | Oncology | Cancer research | Handling missing data | Improving the performance of AI Models | Unstructured: Images | Algorithms can "hallucinate" malignant features in normal synthetic images | Not defined |
| Ali et al. [25] | Gastroenterology | Gastrointestinal endoscopy | 1. Data scarcity due to limited samples<br>2. Avoid data privacy challenges | Developing AI Models | Unstructured: Images | No limitations identified in this paper | Not defined |
| Man et al. [26] | Cardiology | Blood pressure measurement | Data scarcity due to limited samples | Improving the training and evaluation of AI Models | Structured data: Time Series | No limitations identified in this paper | Not defined |
| Ranjbarzadeh et al. [27] | Neurology | Brain tumor segmentation | High Unbalanced datasets | Developing AI Models | Unstructured: Images | GAN technical limitations | Not defined |
| Ali and Shah [28] | Epidemiology | COVID -19 | Data scarcity due to limited samples | Improving the training of AI Models | Unstructured: Images | No limitations identified in this paper | Scoping review |
| He et al. [29] | Neurology | Electroencephalogram classification | 1. Data scarcity due to limited samples<br>2. Avoid overfitting<br>3. High Unbalanced datasets | 1. Developing AI Models<br>2. Improving the performance of AI Models | 1. Unstructured: Images<br>2. Structured data: Time Series<br>3. Structured data: Cross-Sectional | No limitations identified in this paper | Not defined |
| Liu et al. [30] | Oncology | Head and neck cancer | 1. Data scarcity due to limited samples<br>2. High Unbalanced datasets<br>3. Provide complementary information on soft and bone tissues | Improving the performance of AI Models | Unstructured: Images | No limitations identified in this paper | Systematic review and meta-analysis |
| Jiang et al. [31] | Oncology | Computational cytology | 1. Data scarcity due to limited samples<br>2. High Unbalanced datasets | 1. Developing AI Models<br>2. Improving the performance of AI Models | Unstructured: Images | No limitations identified in this paper | Survey |
| Zhu et al. [32] | Endocrinology | Diabetes | Generate population-based datasets to test algorithms in a variety of virtual scenarios, taking into account the high costs and safety issues associated with real clinical trials | Testing AI Models | 1. Structured data: Time Series<br>2. Structured data: Cross-Sectional | No limitations identified in this paper | Systematic review |
| Wen et al. [33] | Oncology | Digital pathology for personalized treatment plans | Challenging data collection | Developing AI Models | Unstructured: Images | No limitations identified in this paper | Not defined |
| Tsilivigkos et al. [34] | Otorhinolaryngology | Application of Deep Learning and imaging | 1. Avoid exposure to ionizing energy and aiding non-experts in diagnosis at the same time<br>2. Eliminate the need for radioactive tracers | Aiding non-experts in diagnosis | Unstructured: Images | No limitations identified in this paper | State-of-the-Art review |
| Lakshmi priya et al. [35] | Gastroenterology | Liver tumour | 1. Huge datasets are required by CNNs<br>2. The use of typical data augmentation techniques (scaling, flipping, rotation, translation, ...) in medical imaging can create distortion of the shape of organs and change the relative position of organs | Improving the performance of AI Models | Unstructured: Images | No limitations identified in this paper | Systematic review |
| Dimitriadis et al. [36] | Oncology | Cancer differentiation (Cancer imaging) | 1. Data scarcity due to limited samples<br>2. High Unbalanced datasets<br>3. Challenging data collection | 1. Improving the performance of AI Models<br>2. Developing AI Models<br>3. Enabling secondary use of data and data sharing | Unstructured: Images | 1. GAN technical limitations<br>2. Due to high heterogeneous, it may not be possible to learn the underlying data properties effectively<br>3. Lack of a standard evaluation benchmark | Systematic review |
| Ben Ali et al. [37] | Cardiology | Interventional Cardiology | 1. Need of improving diagnostics performance<br>2. Try to learn the distribution of the data | 1. Developing AI Models<br>2. Improving the performance of AI Models | 1. Unstructured: Images<br>2. Structured data: Time Series | No limitations identified in this paper | Not defined |
| Ladbury et al. [38] | Oncology | Lung cancer | High Unbalanced datasets | Developing AI Models | Structured: Cross-Sectional | No limitations identified in this paper | Not defined |
| Kruse et al. [39] | Neurology | Diagnose Alzheimer's disease | Improving the performance of diagnostic tools | Developing AI Models | Unstructured: Images | No limitations identified in this paper | Systematic review and meta-analysis |
| Makroum et al. [40] | Endocrinology | Diabetes management | Training and validating ML models | 1. Developing AI Models<br>2. Testing AI Models | Structured data: Time Series | No limitations identified in this paper | Systematic review |
| Balakrishnan et al. [41] | Pneumology | Tuberculosis diagnosis | High Unbalanced datasets | Not specified | Structured data: Cross-Sectional | No limitations identified in this paper | Systematic review |
| Arslan et al. [42] | Oncology | Cancer biology | Testing of different AI Models | Testing AI Models | Structured data: Cross-Sectional | No limitations identified in this paper | Not defined |
| Du et al. [43] | Gynecology | Pregnancy care | Data scarcity due to limited samples | Developing AI Models | Structured data: Cross-Sectional | No limitations identified in this paper | Systematic review |
| Aslam et al. [44] | Neurology | Multiple Sclerosis diagnosis | 1. Data scarcity due to limited samples<br>2. High Unbalanced datasets | Developing AI Models | 1. Unstructured: Images<br>2. Structured data: Cross-Sectional | No limitations identified in this paper | Not defined |
| Zhang and Lindsey [45] | Cardiology | Fetal circulation | Non invasive method of capturing organ-specific and global attributes of fetal circulations | Conducting analysis and studies | 1. Structured data: Time Series<br>2. Unstructured: Videos | Simulated models often assume certain conditions and lose part of the complexity of real-world scenarios | Not defined |
| Ahmed et al. [46] | Endocrinology | Diabetes management | 1. Data scarcity due to limited samples<br>2. High Unbalanced datasets | Developing AI Models | Structured data: Time Series | No limitations identified in this paper | Systematic review |
| Mostapha and Styner [47] | Neurology | Infant brain magnetic resonance imaging for tissue segmentation and disease prediction | High Unbalanced datasets | Developing AI Models | Unstructured: Images | Due to high heterogeneous, it may not be possible to learn the underlying data properties effectively | Not defined |
| Pujante-Otalora et al. [48] | Epidemiology | Computational models for outbreak spread | 1. Simulating realistic scenarios<br>2. Testing models under controlled conditions | 1. Conducting analysis and studies<br>2. Testing AI Models | 1. Structured data: Time Series<br>2. Structured data: Cross-Sectional | No limitations identified in this paper | Systematic review |
| El-Achkar et al. [49] | Nephrology | Kidney citometry | Missing data labels | Not specified | Unstructured: Images | No limitations identified in this paper | Not defined |
| Laubenbacher et al. [50] | Immunology | Mechanistic Digital twins applied to immunology | Capturing the complexity of individual patients and facilitate the development of therapeutic interventions | 1. Conducting analysis and studies<br>2. Optimizing therapies | 1. Unstructured: Images<br>2. Structured data: Time Series<br>3. Structured data: Cross-Sectional | No limitations identified in this paper | Not defined |
| Thakur et al. [51] | Ophthalmology | Glaucoma progression | Improving the utility of TDOCT scans | Conducting analysis and studies | Unstructured: Images | 1. Lack of a standard evaluation benchmark<br>2. GAN technical limitations | Narrative review |
| Gygi et al. [52] | Immunology | Predicting overfitting in immunological applications | Testing of different AI Models | Improving the performance of AI Models | Unstructured: Text | No limitations identified in this paper | Not defined |
| Magalhaes et al. [53] | Gastroenterology | Stomach precancerous lesions | 1. Data scarcity due to limited samples<br>2. High Unbalanced datasets<br>3. Enhancing dataset's quality<br>4. Time-consuming data labeling<br>5. Data anonymization | 1. Enabling secondary use of data and data sharing<br>2. Improving the performance of AI Models | Unstructured: Images | 1. Limited generalization of augmented datasets<br>2. GAN technical limitations<br>3. Synthetic data generators reproduce the bias present in the training data | Not defined |
| Majeed and Zhang [54] | Epidemiology | COVID-19 | 1. Data sharing<br>2. Enhancing dataset's quality<br>3. Privacy preservation<br>4. Data loss prevention<br>5. Improving medical image segmentation, object detection and surgical planning | 1. Developing AI Models<br>2. Improving the performance of AI Models | 1. Unstructured: Text<br>2. Unstructured: Images | No limitations identified in this paper | Not defined |

### 3.3. Specific results

The main findings for each specific topic are detailed in the subsequent subsections.

#### 3.3.1. Main Healthcare Domain and Subdomain

The analysed articles encompass 13 main healthcare domains. These domains, listed from most to least frequently mentioned, are: Oncology (n = 10), Neurology (n = 7), Cardiology (n = 5), Endocrinology (n = 4), Epidemiology (n = 3), Gastroenterology (n = 3), Ophthalmology (n = 3), Immunology (n = 2), Dermatology (n = 1), Gynecology (n = 1), Nephrology (n = 1), Otorhinolaryngology (n = 1), and Pneumology (n = 1). Regarding subdomains, most reviews addressed different subdomains. The only subdomains appearing in more than one review were Diabetes (n = 4) and COVID-19 (n = 2).

#### 3.3.2. Motivations

The reviewed articles identify 45 different motivations for generating synthetic data, which can be consolidated into five main categories: data privacy and security, data scarcity, data quality, AI development, and direct medical and clinical applications. Figure 2 illustrates the distribution of these motivations within each category and their relative frequencies. A detailed table summarising the frequency of these 45 motivations across the articles is provided in Annex 1 of the supplementary material.

**Figure 2.**
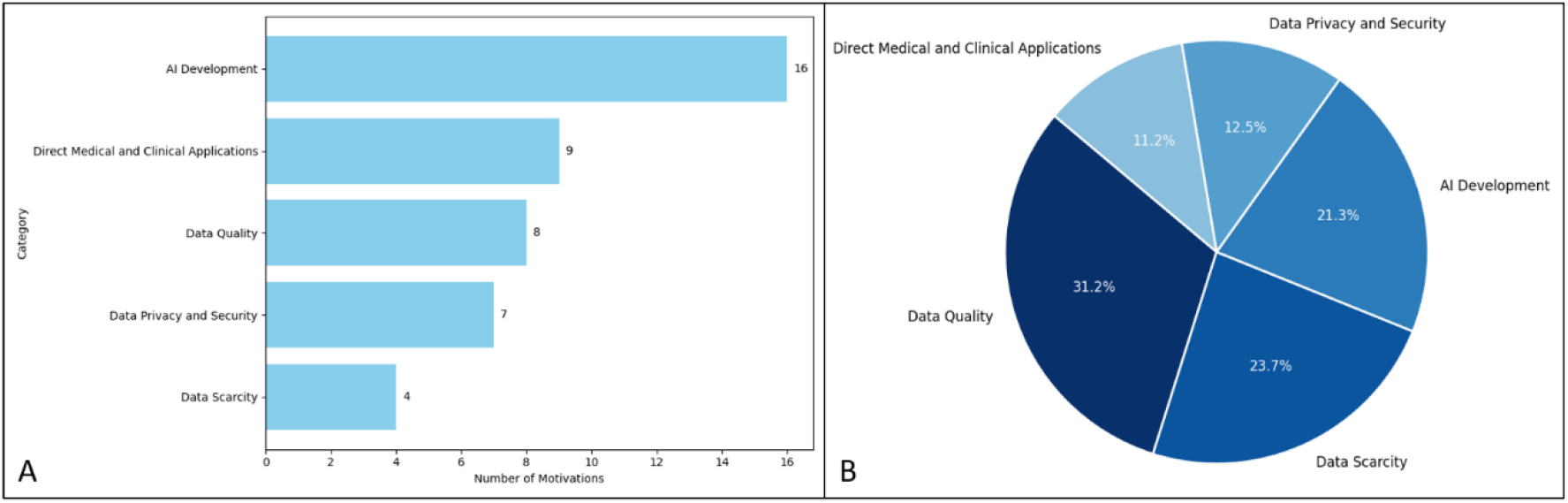
Distribution and Relative Frequency of Motivations Categories. Figure 2A: Distribution of Motivations Across Categories. Figure 2B: Relative Frequency of Category Occurrences.

#### 3.3.3. Future Use

The generated data have been applied in 14 specific use cases, which can be broadly categorized into three main areas: AI development, including training and validation of models and enhancing the generalizability and interpretability of existing models; enabling secondary use of data, such as data sharing and conducting analyses and studies; and enhancing clinical knowledge, serving as educational material or support during diagnosis and therapy. Figure 3 depicts the distribution of these use cases within each category and their relative frequencies. A comprehensive table summarising the occurrences of these use cases in the articles is available in Annex 2 of the supplementary material.

**Figure 3.**
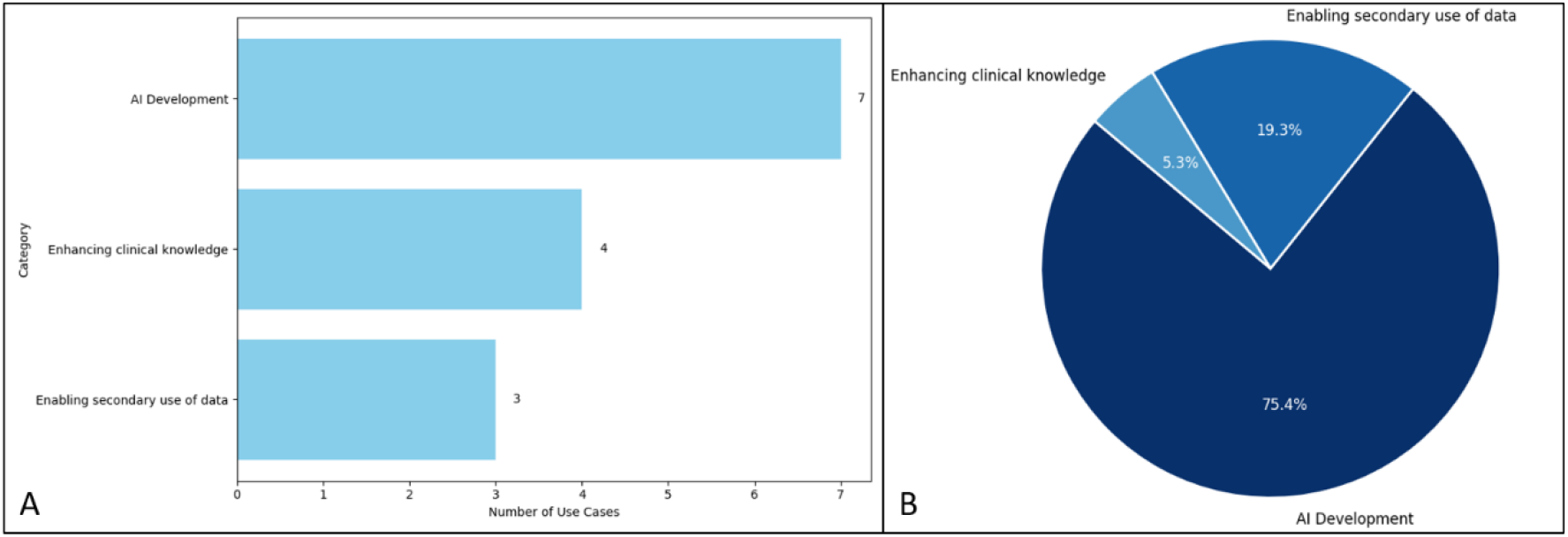
Distribution and Relative Frequency of Use Cases Categories. Figure 3A: Distribution of Use Cases Across Categories. Figure 3B: Relative Frequency of Category Occurrences.

#### 3.3.4. Data Types

Regarding the types of data generated, 30 out of 42 articles (71.43%) refer to the generation of unstructured data, while 18 out of 42 articles (42.86%) refer to the generation of structured data. Among the unstructured data, images are the most commonly generated type (n = 28), followed by text (n = 2) and video (n = 1). For structured data, both time series (n = 11) and cross-sectional data (data collected at a single point in time, providing a snapshot) (n = 12) are similarly represented.

#### 3.3.5. Limitations

Only 14 of the 42 reviews mentioned limitations. The most frequently cited limitations were related to GAN technical issues (n = 7), the absence of a standard evaluation benchmark (n = 5), and the transmission of biases from training data to synthetic data (n = 5). Additional limitations included the challenges posed by small and heterogeneous datasets, issues with the generalization of augmented data, and the loss of complexity inherent in real-world scenarios.

#### 3.3.6. Review Type

Among the 42 reviews analysed, 22 do not specify their type. The remaining 20 reviews are categorized as follows: 10 Systematic reviews, 3 Systematic reviews and Meta-analyses, 2 Scoping reviews, 2 Narrative reviews, 2 Surveys, and 1 State of the Art review.

## 4. Discussion

In this section, we emphasize the contributions of this scoping review of reviews. This study has systematically analysed all the reviews from the last decade that discuss the generation of synthetic data in different domains of healthcare. Specifically, we examined the main domains and subdomains where synthetic data are being generated, the motivations behind this generation, the potential future applications of these data, the types of data generated, the limitations identified by the authors regarding synthetic data generation, and the types of reviews analysed. Understanding the current state and potential growth areas for synthetic data in healthcare is important to identify how these data can help in addressing key challenges and contribute to advancements in healthcare domain. General reflections from the full-text screening process suggest that while synthetic data generation is a promising field with significant potential to address many current challenges in a multidisciplinary manner, it still needs to be further integrated and applied across the different areas of healthcare. This highlights the need for focused research on the current generation and applications of synthetic data to fully realize its potential and overcome existing barriers to its widespread adoption.

Our findings towards healthcare domains indicate that synthetic data generation is most prevalent in fields such as Oncology, Neurology, and Cardiology, which reflects a high demand for data in these areas due to challenges like data scarcity and privacy concerns. Less frequently mentioned domains, including Dermatology, Gynecology, and Pneumology, suggest emerging interest and potential for further exploration. Regarding subdomains, there is greater variety, as most articles do not share common subdomains. The exceptions are the endocrinology articles focusing on diabetes [22,32,40,46] and articles addressing COVID-19 [28,54], reflecting the impact of these issues on society. This wide range of healthcare domains and subdomains where synthetic data is currently generated illustrates the versatility of this technology in the healthcare sector.

Arising from our analyses, we have found that the motivations for generating synthetic data, while diverse, raise several critical concerns that are worthy of further consideration. These motivations can be broadly classified into five categories: data privacy and security, data scarcity, data quality, AI development, and direct medical and clinical applications. The emphasis on data privacy and security, while valid, often oversimplifies the complexities involved in ensuring truly anonymised synthetic datasets. The assumption that synthetic data can completely mitigate privacy risks overlooks potential vulnerabilities in the data generation process, such as re-identification risks if synthetic data are not sufficiently differentiated from real data especially in the health field. The issue of data scarcity, especially in cases of rare diseases in patients, highlights an important gap in health research that allows synthetic data to provide an answer. However, the reliance on synthetic data to fill these gaps can lead to a false sense of data adequacy and we must consider principles mentioned above to ensure a minimum of quality. This limitation is often exacerbated by the quality of the available data, which is often unbalanced or incomplete.

The development of AI as a motivation for synthetic data generation is compelling, given the need for large datasets to train sophisticated models. However, the quality of the AI models produced is intrinsically linked to the quality of the synthetic data. The risk of perpetuating existing biases or introducing new ones is a major concern that needs to be rigorously addressed. Potential pitfalls, such as privacy risks, data quality issues and biases, make it clear that the recent AI Act regulation adopted by the European Commission, and which entered into force on 1 August 2024 will need to consider address these issues.

Furthermore, synthetic data are used to promote open science and secondary data use through data sharing for analyses and studies, as well as to improve clinical knowledge by assisting in various tasks such as diagnostics and personalized therapies or serving as educational material. This demonstrates that synthetic data are valuable not only for technological advancements in the healthcare sector, such as the development of decision support systems, but also in the scientific and academic fields, facilitating open research and the sharing of higher quality information.

The reviewed articles predominantly discuss the generation of unstructured data, particularly images, reflecting the critical role of medical imaging in healthcare. However, there is a notable gap in the generation of other types of unstructured data, such as video and text, which are increasingly relevant with the advent of more complex generative models. Structured data, including time series and cross-sectional data, also play a significant role in capturing comprehensive patient information and warrant further investigation to enhance their utility in healthcare applications.

Only one-third of the articles identified limitations related to synthetic data generation. The most frequently mentioned limitation was technical issues with GAN models, such as instability during training and mode collapse. Additionally, there is a strong emphasis on the need for standard evaluation benchmarks, as highlighted in other studies like the one conducted by Murtaza et al. [10], and the transmission of biases present in the original data to the synthetic data, which is a necessary consideration. These three limitations are not exclusive to the health domain but are relevant to any type of synthetic data. This implies that as research on synthetic data generation advances, its application across all the different domains will continue to evolve.

Lastly, it is worth noting that several studies have identified synthetic data generators for specific domains that are widely accepted and commonly used within the community. This practice should become more widespread in the coming years, fostering scientific progress across various health sectors and promoting research in synthetic data. The synthetic data generators referenced in several studies include the UVA/PADOVA Type 1 Diabetes Simulator [55], which is cited by 3 out of the 4 articles concerning diabetes; and the simulator for single-cell RNA sequencing data (SPLATTER) [56], which is cited by the article focused on cancer biology. These two synthetic data generators serve as an ideal reference for the scientific community, facilitating greater access to quality health data without compromising individual privacy.

### 4.1. Limitations and Strengths of the study

This scoping review demonstrates several strengths. Firstly, the exclusive analysis of domain-specific reviews ensures that the extracted content is reliable and relevant within the specified domain. Additionally, a systematic approach was employed, adhering to established guidelines and standards, enhancing the rigor of the review. To minimize bias, the extraction and analysis of results were conducted by three different researchers. These combined strengths ensure that the information provided in this review is aligned with the current knowledge in the literature.

However, a notable limitation of this study is the rapid growth of AI and its scientific output, which means that this review provides only a baseline snapshot that will continue to evolve in the coming years. Furthermore, the exclusion of articles discussing the future potential of synthetic data might have limited the scope of insights into emerging trends and anticipated developments in this field. Despite these limitations, this Scoping review complements the more technologically focused reviews by providing insights into the clinical and practical applications of synthetic data generation within healthcare, offering a robust foundation for future research in this field.

## 5. Conclusions and Future Work

Synthetic data generation is a promising technology with high potential to enhance healthcare and healthcare research. This study has reviewed 42 articles to provide a comprehensive overview of the primary healthcare domains and subdomains where these techniques are applied, their motivations, the types of data generated, their future uses, and the limitations encountered. While the analysis indicates that synthetic data with various characteristics and typologies are currently being generated across many main healthcare domains, the technology’s versatility and relatively early stage suggest considerable potential for future applications.

Firstly, further investigation and application are needed in domains where synthetic data is just beginning to be used, such as Immunology, Dermatology, and Gynecology. It is also essential to extend their application to new domains and subdomains that have not adopted these techniques yet. Additionally, the generation of videos and text, which remains underexplored in the healthcare field, has great possibilities, especially with the recent advancements in generative models and Large Language Models. Finally, efforts are needed to define a standard evaluation benchmark, as previously highlighted in the literature, and to develop reference models for specific domains such as the UVA/PADOVA Type 1 Diabetes Simulator to foster open research and investigation in the generation of synthetic health data.

## Data Availability

All data produced in the present work are contained in the manuscript

## Declaration of generative AI and AI-assisted technologies in the writing process

During the preparation of this work, the authors used ChatGPT 4 in order to proof of read some of the text. After using this tool/service, the authors reviewed and edited the content as needed and take full responsibility for the content of the published article.

