## Supplementary material for "Synthetic Data Generation in Healthcare: A Scoping Review of reviews on domains, motivations, and future applications"

### Supplementary material – Annex 1

*TABLE 1. Motivation details*

| Motivations | Frequency |
|---|---|
| Avoid data privacy challenges | 4 |
| Avoid exposure to ionizing energy and aiding non-experts in diagnosis at the same time. | 1 |
| Avoid overfitting | 1 |
| Avoid time consuming data collection and labelling. | 1 |
| Building a benchmark database for evaluation purposes | 1 |
| Capturing the complexity of individual patients and facilitate the development of therapeutic interventions | 1 |
| Challenging data collection | 2 |
| Data anonymization | 1 |
| Data 1os prevention | 1 |
| Data scarcity due to limited samples (low disease prevalence) | 15 |
| Data sharing | 1 |
| De-identification | 1 |
| Difficulty in collecting large consented datasets of highly vulnerable patients under demanding care plans ( Data sharing and privacy). | 1 |
| Disease simulation and surgery planning | 1 |
| Eliminate the need for radioactive tracers | 1 |
| Enable comparison between different methods | 1 |
| Enhancing dataset’s quality | 2 |
| Generate population-based datasets to test algorithms in a variety of virtual scenarios, taking into account the high costs and safety issues associated with real clinical trials. | 1 |
| Handling missing data | 2 |
| High heterogeneity between tumours | 1 |
| High Unbalanced datasets | 15 |
| Huge datasets are required by CNNs | 1 |
| Improving cancer detection, diagnosis, tumour profiling, treatment planning and monitoring | 1 |
| Improving medical image segmentation, object detection and surgical planning | 1 |
| Improving ML Systems | 1 |
| Improving the performance of diagnostic tools | 1 |
| Improving the quality of images | 1 |
| Improving the quality of the data | 1 |
| Improving the utility of TDOCT scans | 1 |
| Lack of large annotated databases available | 1 |
| Low data accessibility due to regulatory frameworks, ethics and privacy (GDPR) | 1 |
| Low data quality | 1 |
| Missing data annotation | 1 |
| Missing data labels | 2 |
| Need of improving diagnostics performance | 1 |
| Non invasive method of capturing organ-specific and global attributes of fetal circulations | 1 |
| Privacy preservation | 1 |
| Provide complementary information on soft and bone tissues. | 1 |
| Simulating realistic scenarios | 1 |
| Testing models under controlled conditions | 1 |
| Testing of different AI Models | 2 |
| The use of typical data augmentation techniques (scaling, flipping, rotation, translation, …) in medical imaging can create distortion of the shape of organs and change the relative position of organs. | 1 |
| Time-consuming data labeling | 1 |
| Training and validating ML models | 1 |
| Try to learn about the distribution of the data | 1 |

### Supplementary material - Annex 2

*TABLE 2. Future uses details*

| Future Use | Frequency |
|---|---|
| Aiding non-experts in diagnosis | 1 |
| Conducting analysis and studies | 6 |
| Data sharing | 1 |
| Developing AI Models | 22 |
| Developing Large-scale Models | 1 |
| Enabling secondary use of data and data sharing | 4 |
| Improving the generalizability and interpretability of AI Models | 1 |
| Improving the performance of AI Models | 12 |
| Improving the training and evaluation of AI Models | 1 |
| Improving the training of AI Models | 1 |
| Medical education | 1 |
| Not specified | 3 |
| Optimizating therapies | 1 |
| Testing AI Models | 5 |
